# Protocol for a randomized controlled trial testing inhaled nitric oxide therapy in spontaneously breathing patients with COVID-19

**DOI:** 10.1101/2020.03.10.20033522

**Authors:** Chong Lei, Binxiao Su, Hailong Dong, Bijan Safaee Fakhr, Luigi Giuseppe Grassi, Raffaele Di Fenza, Stefano Gianni, Riccardo Pinciroli, Emanuele Vassena, Caio Cesar Araujo Morais, Andrea Bellavia, Stefano Spina, Robert Kacmarek, Lorenzo Berra

**Affiliations:** Department of Anesthesiology and Perioperative Medicine, Xijing Hospital, the Fourth Military Medical University. Xi’an, Shaanxi, China; Intensive Care Unit, Department of Anesthesiology and Perioperative Medicine, Xijing Hospital, the Fourth Military Medical University. Xi’an, Shaanxi, China; Department of Anesthesia, Critical Care and Pain Medicine, Massachusetts General Hospital, Boston, Massachusetts, USA; Department of Environmental Health, Harvard T.H. Chan School of Public Health, Boston, Massachusetts, USA

## Abstract

**Introduction:** the current worldwide outbreak of Coronavirus disease 2019 (COVID-19) due to a novel coronavirus (SARS-CoV-2) is seriously threatening the public health. The number of infected patients is continuously increasing and the need for Intensive Care Unit admission ranges from 5 to 26%. The mortality is reported to be around 3.4% with higher values for the elderly and in patients with comorbidities. Moreover, this condition is challenging the healthcare system where the outbreak reached its highest value. To date there is still no available treatment for SARS-CoV-2. Clinical and preclinical evidence suggests that nitric oxide (NO) has a beneficial effect on the coronavirus-mediated acute respiratory syndrome, and this can be related to its viricidal effect. The time from the symptoms’ onset to the development of severe respiratory distress is relatively long. We hypothesize that high concentrations of inhaled NO administered during early phases of COVID-19 infection can prevent the progression of the disease.

**Methods and analysis:** This is a multicenter randomized controlled trial. Spontaneous breathing patients admitted to the hospital for symptomatic COVID-19 infection will be eligible to enter the study. Patients in the treatment group will receive inhaled NO at high doses (140-180 parts per million) for 30 minutes, 2 sessions every day for 14 days in addition to the hospital care. Patient in the control group will receive only hospital care. The primary outcome is the percentage of patients requiring endotracheal intubation due to the progression of the disease in the first 28 days from enrollment in the study. Secondary outcomes include mortality at 28 days, proportion of negative test for SARS-CoV-2 at 7 days and time to clinical recovery.

**Ethics and dissemination:** The trial protocol has been approved at the Investigation Review Boards of Xijing Hospital (Xi’an, China) and The Partners Human Research Committee of Massachusetts General Hospital (Boston, USA) is pending. Recruitment is expected to start in March 2020. Results of this study will be published in scientific journals, presented at scientific meetings, and on related website or media in fighting this widespread contagious disease.

**Trial registration:** Clinicaltrials.gov. NCT submitted

## Introduction

The novel Coronavirus 2019 (SARS-CoV-2) has been first identified in China in December 2019 [1]. As of today, more than 110,000 cases have been reported (with more than 4,000 deaths) and, while the majority of them is still concentrated in China, the infection has spread across the country’s border to many Asian countries, as well as to Europe and to the USA [2].

SARS-CoV-2 is a positive single-stranded RNA virus belonging to the family *Coronaviridae*. The current outbreak would result from the acquired ability of the virus to undergo human to human transmission [3], after the jump from the original animal reservoir (most likely a bat).

In the human host, SARS-CoV-2 causes a respiratory syndrome (named COVID-19) which can range from a mild involvement of the upper airways to a severe pneumonia with Acute Respiratory Syndrome (ARS) and the need of mechanical ventilation in an intensive care unit (ICU). In early case series, the risk of critical care admission ranges between 5 and 26%, with higher values for the elderly and in patients with comorbidities or chronic diseases [4,5]. The time from symptoms onset to development of severe respiratory distress is relatively long, with a median time of 8 days in which the patient remains in a state of mild disease characterized by dry cough, fever and mild hypoxia requiring oxygen supplementation. Once severe disease with the necessity of ICU develops, the outcome worsens decisively, with mortality raising from 3.4% to 61% [4,6]. Moreover, ICU staying poses a serious strain on resource-limited hospitals. Thus, preventing disease progression during the mild phase would be highly desirable both in terms of morbidity and mortality improvement and healthcare resource-sparing. At the present time, there are no proven etiological treatments for COVID-19 but some ongoing clinical trials are testing the effects of anti-viral drugs (NCT04252664 on ClinicalTrials.gov).

In 2004, a pilot study showed that low dose inhaled Nitric Oxide (NO) was able to shorten the time of ventilatory support during the SARS epidemic, sustained by another coronavirus, SARS-CoV [7]. In an in vitro study, the NO donor compound S-nitroso-N-acteylpenicillamine increased the survival rate in an in vitro model of SARS-CoV infected Monkey’s epithelial cells [8], suggesting a possible viricidal effect of the gas. Inhaled NO at high doses can be administered safely and is known for potential microbicidal effects [9–12]. While further in-vitro testing for this specific virus is recommended, we propose a randomized clinical trial to test the effectiveness of inhaled NO in preventing progression of COVID-19 disease, when administered at an early stage.

## Methods and analysis

### Study design

Multicenter randomized clinical trial.

### Study setting

The coordinating center for China of this study is Xijing Hospital, the Fourth Military Medical University in Xi’an, Shaanxi, China.

The coordinating center for other centers is the Massachusetts General Hospital in Boston, Massachusetts, USA. This study is open to other centers willing to participate.

For more information see section “Contacts”.

### Eligibility criteria

Adults in-hospital patients (≥18 years old) will be recruited within 72 hours after confirmation of COVID-19 by real-time Reverse Transcriptase Polymerase Chain Reaction (RT-PCR) on oropharyngeal or nasopharyngeal swabs or stool samples. Diagnosis can also be obtained by detection of COVID-19 IgM/IgG antibodies in serum, plasma or whole blood sample.

Symptoms of COVID-19 must include presence of fever of at least 36.6 °C from the axillary site or 37.2 °C from the oral site or 37.6 °C from the rectal / tympanic site.

In addition, patient must be spontaneously breathing with a respiratory rate of at least 24 breaths per minute and/or an objective persistent cough consistent with COVID-19 symptoms.

The patients with or without hypoxia will be included. Gas exchange and ventilation may be assisted by means of any continuous positive airway pressure (CPAP) system, or any system of non-invasive ventilation (NIV) with positive end-expiratory pressure (PEEP) ≤ 10 cmH_2_O.

Criteria of exclusion are pregnancy (all women in fertile age should be tested for pregnancy before being enrolled); presence of an open tracheostomy; therapy with high-flow nasal cannula; clinical contraindication to NO gas delivery, as judged by the attending physician; hospitalized and confirmed diagnosis of COVID-19 for more than 72 hours.

Table 1 summarize inclusion and exclusion criteria.

**Table 1.**
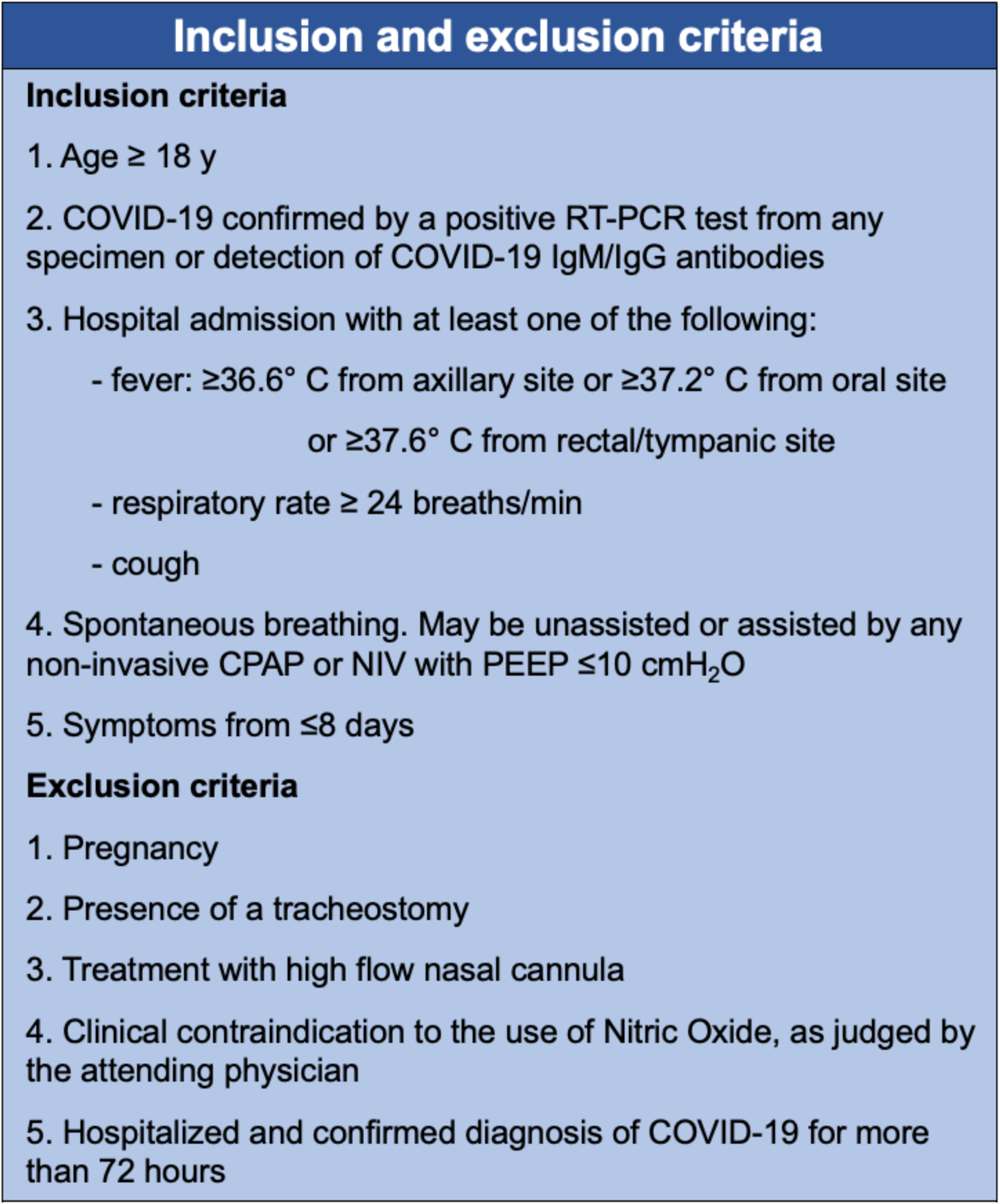
RT-PCR= Reverse Transcription-Polymerase Chain Reaction. CPAP= Continuous Positive Airway Pressure. NIV= Non-Invasive Ventilation. PEEP= Positive End Expiratory Pressure

### Interventions

This is a randomized (1:1) controlled, parallel arm clinical trial to verify that brief periods of inhaled NO at high doses can improve clinical course reducing the number of patients who undergo endotracheal intubation for COVID-19 in the first 28 days after enrollment in the study (primary endpoint).

Once admitted to the hospital with a confirmed diagnosis of COVID-19 (obtained by RT-PCR for SARS-COVID-2 virus, or positive in COVID-19 IgM/IgG antibodies) the patients will be screened. Eligible patients randomized to the treatment group will receive iNO to target an average inspiratory concentration between 140 to 180 ppm. The administration of iNO will be performed through a non-invasive ventilation circuit with a range between 2 and 10 cmH_2_O of positive end expiratory pressure (PEEP) or a non-rebreathing mask without positive end expiratory pressure depending on the clinical needs of the patient and availability of respiratory equipment. The patients will undergo 2 sessions per day with the duration between 20 and 30 minutes. A representation of the administration setting in shown in Figure 1.

**Figure 1.**
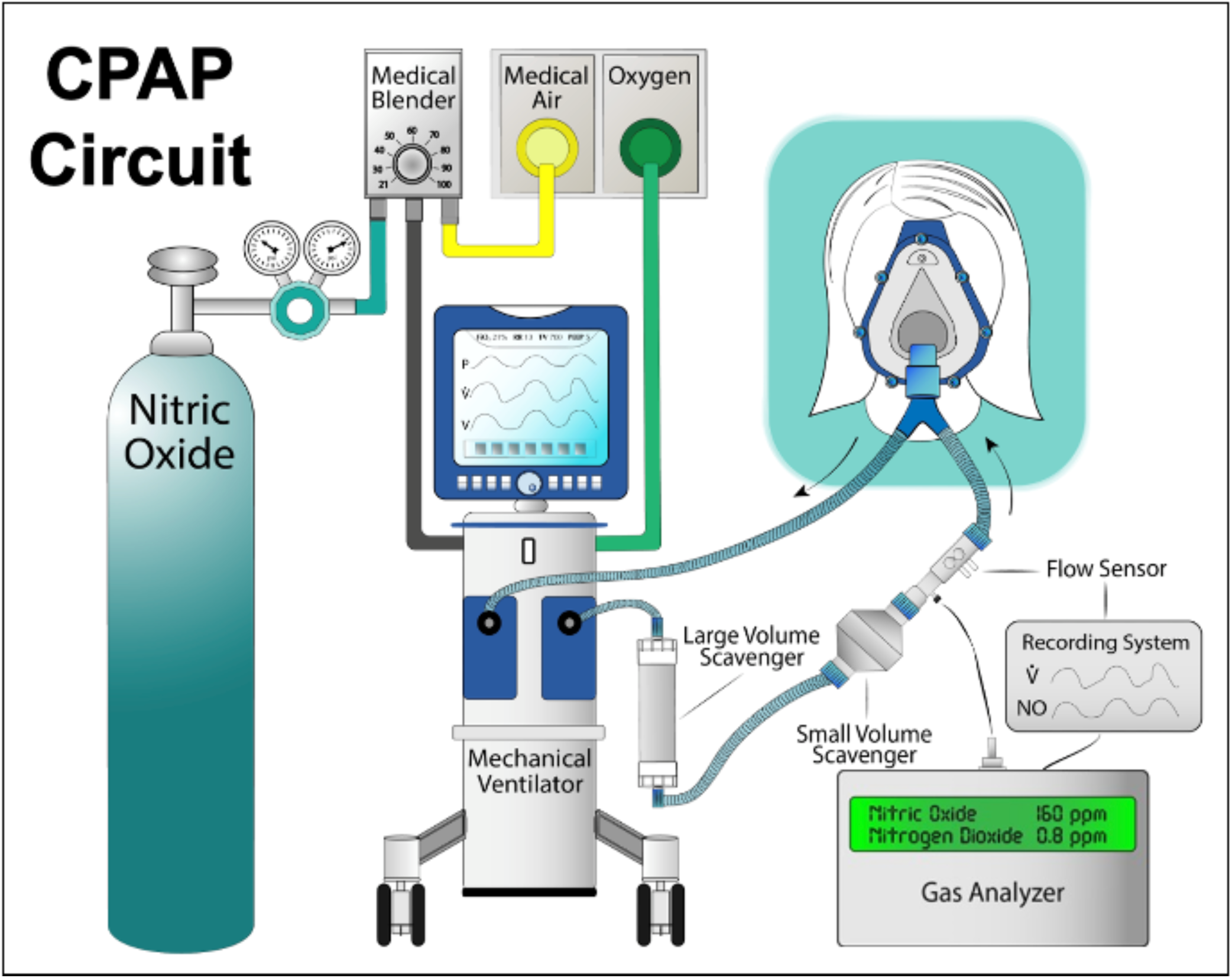
A schematic example of the delivery system mounted on commonly available ventilator

The NO gas therapy will continue for 14 days or until the primary outcome event is reached (i.e., clinical deterioration with indication for intubation and mechanical ventilation). Other criteria for the interruption of the therapy are: negative result on RT-PCR for the presence of the coronavirus (SARS-CoV-2, endpoint 3); hospital discharge or recovery from the symptoms, defined as normalization of fever, respiratory rate, and alleviation of cough, sustained for at least 72 hours (endpoint 4). A flowchart of the study is presented in Figure 2.

**Figure 2.**
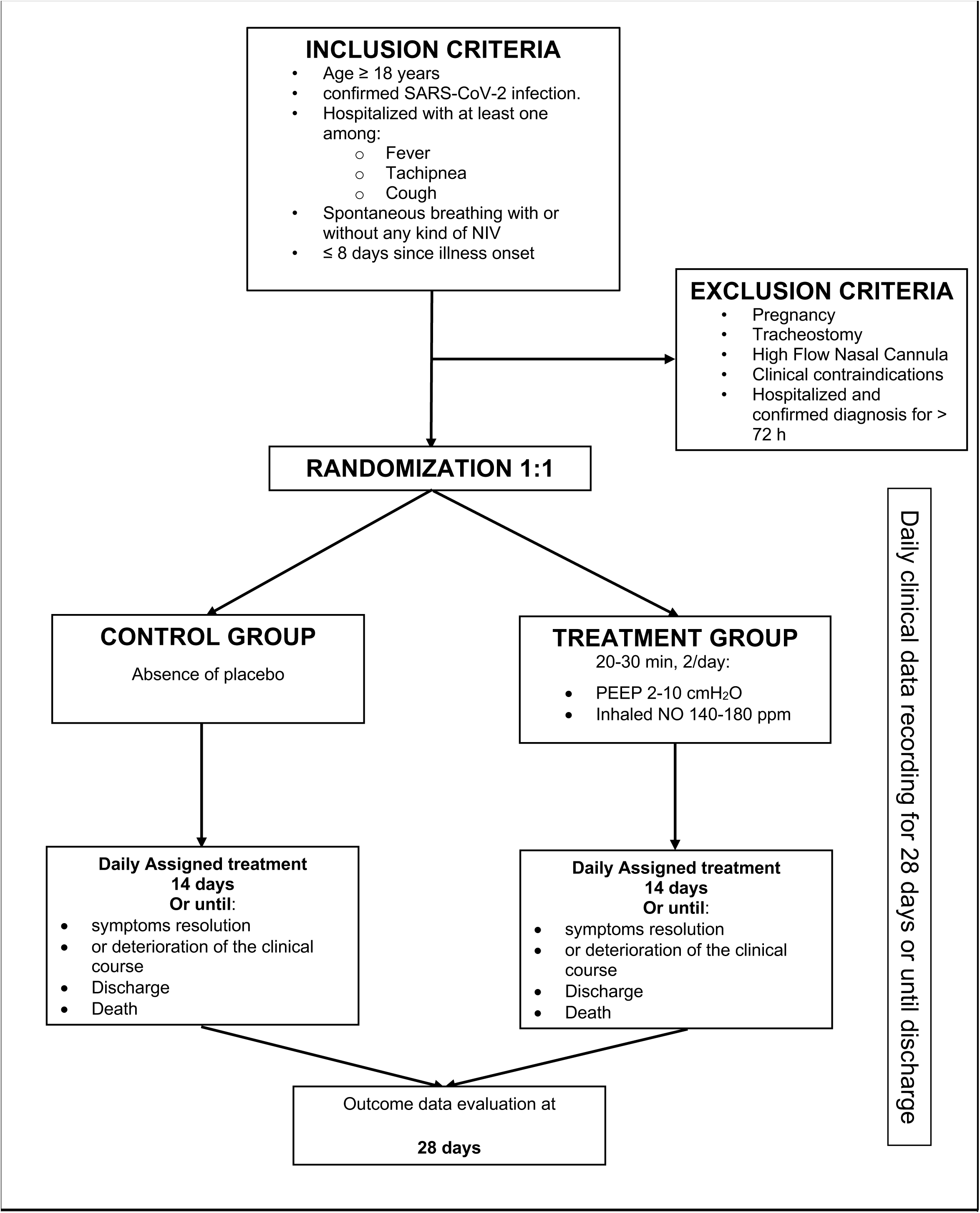
Study Flowchart.

The evaluation of the patients will continue for 28 day after the enrollment. Since treatment with inhaled NO can lead to increase in plasmatic methemoglobin, the blood levels of methemoglobin will be monitored via non-invasive CO-oximeter or methemoglobin levels in blood. If methemoglobin levels rise above 5% at any point of the study, inhaled NO concentration will be immediately stopped until the subsequent dose. For safety reason we will also evaluate NO_2_ level in order to keep it below 5 ppm. If we observe an increase above this value, we will immediately interrupt the administration of NO until the subsequent dose.

Patients assigned to the control group will not receive any gas therapy.

### Outcomes

The primary outcome will be the proportion of patients who progress to a severe form of the disease, defined as the indication given by the attending physician to intubation and mechanical ventilation in the first 28 days after enrollment in the study. Patients in severe respiratory failure and/or ventilatory distress with indication to intubation but concomitant DNI (Do Not Intubate) will meet criteria for primary endpoint. The proportion of intubation will be calculated at 28 days from study enrollment.

Secondary outcomes include:

- Mortality from any cause at 28 days (endpoint 2)
- Proportion of negative conversion of SARS-CoV-2 from upper respiratory tract specimens at 7 days (endpoint 3)
- Time to clinical recovery, defined as: normalization of fever (≤ 36.6 C from the axillary site; or ≤ 37.2 C from the oral site; or ≤ 37.8 C from the rectal/tympanic site) plus normalization of respiratory rate < 24 breaths/min and alleviation of cough (defined as mild or absent in a patient-reported scale of severe>>moderate>>mild>>absent) or discharge home (endpoint 4).

### Data collection

Clinical information including medical history and laboratory exams will be obtained from the medical charts and prospectively recorded until discharge or death. Collection of study variables will be managed by the outcome assessors by using a dedicated patient’s file on Studytrax. The 28 days follow up, if the patient is discharged from the hospital, will be performed by a phone call.

Outcome assessors, treatment providers and the principal investigator will obtain unique usernames and password to transfer all data to a Studytrax page dedicated to the study. Data access is restricted only to authorized member of the team which will be responsible for strict confidentiality all times. The signed informed consent will be kept in a secure place for at least 5 years after study completion.

### Sample size calculation

Based on previously published data on COVID 19 [4], we predict an incidence of intubation and mechanical ventilation of 12.3%. However, because of the novel nature of this study, we do not have enough data to predict the effect of inhaled NO on the primary outcome. The dynamic trend of this ongoing outbreak also prevents us to determine the rate of enrollment that we can achieve. As such, in Table 2 we are displaying the sample size calculation for our primary outcome, for an alpha level of 0.05 and a power level of 0.8, under different scenario. We will perform a first analysis based on 240 patients, evaluating the actual incidence of the primary outcome, effect of the treatment and the actual rate of enrollment. The PI will decide, together with the DSMB, whether to continue enrollment to 340 patients, if a sufficient rate of enrollment and power had not been achieved with the original sample size. Based on results reported in Table 2, the same step could then be repeated increasing sample size from 340 to 500 patients, from 500 patients to 760 and from 760 patients to 1260.

**Table 2.**
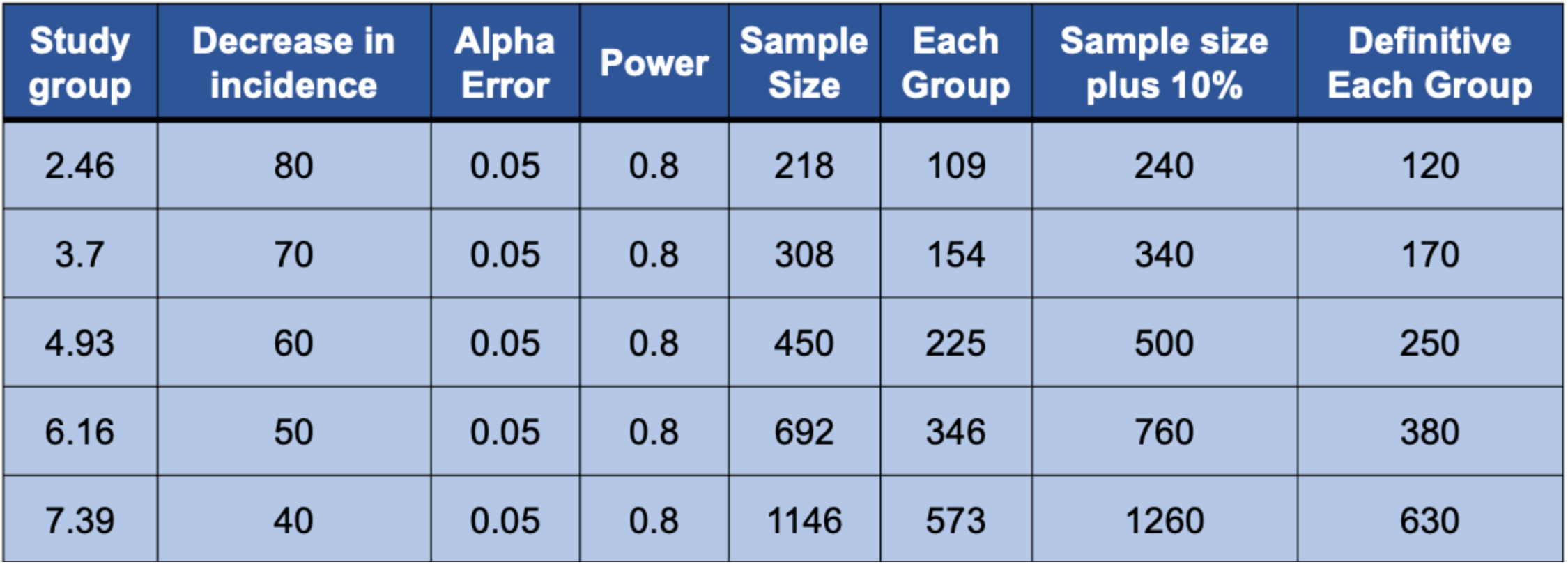
Sample size calculation for any given decrease in incidence of intubation induced by NO

### Statistical analysis

Data will be analyzed following the intention to treat analysis principle. Demographic and clinical data will be presented as proportions for categorical outcomes and mean plus standard deviation or median plus interquartile range for continuous outcomes. Comparison between groups will be made with the X^2^ test or the Fisher exact test for categorical variables and with the t-test or Wilcoxon rank-sum test for continuous variables. For the t-test the normality distribution will be evaluated.

Time to event outcomes will be analyzed by estimating survival curves over treated and non-treated using the Kaplan-Meier method.

A subgroup analysis will be performed with multiple linear regression, as well as logistic and cox regression models to adjust for age, pulmonary comorbidities and co-administration of other experimental treatment.

### Data monitoring

Data will be monitored by the principal investigator (PI) in collaboration with an independent Data and Safety Monitoring Board (DSMB). PI, and DSMB will monitor adverse events and quality of the data and provide recommendations. The PI will monitor compliance to safety rules every 20 patients. Every violation will be reported to the DSMB. Peripheral centers will be provided with a data sheet providing safety rules and will be instructed to report every violation to the coordinating hospital. Before the beginning of the study, the DSMB will meet to decide safety rules and stopping guidelines.

We will not perform any interim analysis, but the DSMB will be granted the authority to stop the trial at any point due to safety concerns.

### Conclusion

The aim of this trial is to evaluate whether high dose of NO administered at an early stage can safely reduce or prevent the progression of COVID-19 disease. This study is open to every center that wants to participate. If interested in participating in the study, please contact the Principal Investigator.

## Data Availability

This is a trial design for an upcoming study on COVID-19. We hypothesize that high concentrations of inhaled NO administered during early phases of COVID-19 infection can prevent the progression of the disease.

## Contacts

Lorenzo Berra, MD

Bijan Safaee Fakhr, MD

Chong LEI, MD and PhD

## Authors’ contributions

Authorship for this trial will be given to key personnel involved in trial design, personnel training, recruitment, data collection, statistical plan and data analysis. There are no publication restrictions. CL, BS, HD, LB, RP were responsible for conceptualizing trial design. CL and LB managed patient safety protocol. CL, BS, HD, LB, BSF, RDF are responsible for recruitment, enrolment and data collection. AB, BSF, LG are responsible for power calculation, statistical plan and data analysis. CL, BS, HD, LB, EV, BSF, RDF, SG and CCAM trained personnel for the clinical trial and built systems for nitric oxide delivery and monitoring. All authors have critically revised the study protocol and approved the final version. All authors agree to be accountable for the accuracy and integrity of all aspects of this trial.

## Funding statement

Local departmental funds

## Competing interests’ statement

LB salaries are partially supported by NIH/NHLBI 1 K23 HL128882-01A1.

## Notes

### Competing Interest Statement

Lorenzo Berra salaries are partially supported by NIH/NHLBI 1 K23 HL128882-01A1.

